# Association of sepsis and delayed cerebral ischemia in patients with aneurysmal subarachnoid hemorrhage

**DOI:** 10.1101/2024.01.27.24301874

**Authors:** Franz-Simon Centner, Holger Wenz, Mariella Eliana Oster, Franz-Joseph Dally, Johannes Sauter-Servaes, Tanja Pelzer, Jochen Johannes Schoettler, Bianka Hahn, Amr Abdulazim, Katharina Antonia Margarete Hackenberg, Christoph Groden, Joerg Krebs, Manfred Thiel, Nima Etminan, Máté Elod Maros

## Abstract

**Background:** Although sepsis and delayed cerebral ischemia (DCI) are severe complications in patients with aneurysmal subarachnoid hemorrhage (aSAH) and share pathophysiological features, their interrelation and additive effect on functional outcome is uncertain. We investigated the association of sepsis with DCI and their cumulative effect on functional outcome in patients with aSAH using current sepsis-3 definition.

**Methods:** Patients admitted to our hospital between 11/2014-11/2018 for aSAH were retrospectively analyzed. The main explanatory variable was sepsis, diagnosed using sepsis-3 criteria. Endpoints were DCI and functional outcome at hospital discharge (modified Rankin Scale (mRS) 0-3 vs. 4-6). Propensity score matching (PSM) and multivariable logistic regressions were performed.

**Results:** Of 238 patients with aSAH, 55 (23%) developed sepsis and 74 (31%) DCI. After PSM, aSAH patients with sepsis displayed significantly worse functional outcome (p<0.01) and longer ICU stay (p=0.046). Sepsis was independently associated with DCI (OR=2.46, 95%CI: 1.28-4.72, p<0.01). However, after exclusion of patients who developed sepsis before (OR=1.59, 95%CI: 0.78-3.24, p=0.21) or after DCI (OR=0.85, 95%CI: 0.37-1.95, p=0.70) this statistical association did not remain. Good functional outcome gradually decreased from 56% (76/135) in patients with neither sepsis nor DCI, to 43% (21/48) in those with no sepsis but DCI, to 34% (10/29) with sepsis but no DCI and to 8% (2/26) in patients with both sepsis and DCI.

**Conclusions:** Our study demonstrates a strong association between sepsis, DCI and functional outcome in patients with aSAH and suggests a complex interplay resulting in a cumulative effect towards poor functional outcome, which warrants further studies.

## Introduction

Both sepsis and delayed cerebral ischemia (DCI) are severe complications in patients with aneurysmal subarachnoid hemorrhage (aSAH) and are significant determinants for poor outcome^1–4^. The term DCI presents a pro-ischemic complication in patients who survive the initial aSAH ictus^5,6^. At the same time sepsis can also result in acute and long-term brain damage via cerebral ischemia^7,8^. Both sepsis and DCI are characterized by a dysregulation in vascular regulation and integrity^9,10^. DCI was mainly thought to be caused by cerebral vasospasm, however, recent studies support the concept of a multifactorial pathophysiology, including microcirculatory disturbances due to microthrombosis and neuroinflammation^4,11^. Microcirculatory dysfunction is also a key mechanism of septic organ dysfunction caused by direct effects of circulating mediators, capillary leak and microthrombus formation, which can affect any organ including the brain^7,10^. In this context, it was recently shown that systemic infection can amplify intracranial inflammatory responses in patients with aSAH^12^.

Considering these overlapping pathophysiological features, we hypothesized that in aSAH patients the development of sepsis and DCI might be associated and may have an additive effect resulting in poor functional outcome. However, current data on the relation of sepsis and DCI in patients with aSAH are limited and inconclusive^13^. Although nosocomial infection was independently associated with DCI in an exploratory analysis^13^, two preceding studies did not find an association between sepsis and DCI^9,14^. These studies were, however, performed before 2016^9,14^ and did not use the updated sepsis definition (sepsis-3), which focuses on septic organ dysfunction^15^. Therefore, we investigated the association of sepsis with DCI and a potential cumulative effect on functional outcome in aSAH patients using current sepsis definition.

## Methods

### Study design and setting, patient selection

This single-center retrospective cohort study was conducted at the 25-bed intensive care unit (ICU) of the Department of Anaesthesiology, Surgical Intensive Care Medicine and Pain Medicine at the University Medical Center Mannheim, Germany, where aSAH patients were treated and managed in close collaboration with the Department of Neurosurgery. The Medical Ethics Commission II of Medical Faculty Mannheim, University of Heidelberg, approved the study design as well as reanalysis of neuroradiological reports and imaging data (reference nr.: 2019-1096R, 2017-825R-MA and 2017-828R-MA; date of approval: 01/10/2019). The need for informed consent was waived because of the retrospective nature of the study. Study design and reporting adhered to the STROBE guidelines (PMID: 18064739, STrengthening the Reporting of OBservational studies in Epidemiology, http://www.strobe-statement.org). The study was registered at the German Clinical Trials Register (ID: DRKS00030748, https://drks.de/search/en/trial/DRKS00030748).

Data on all patients (age ≥ 18 years) treated in our ICU between 11/2014 and 11/2018 were extracted from an electronic health record database (Fig. 1). Of 3961 patients, 340 (8.6%) patients were preselected via ICD codes for spontaneous subarachnoid hemorrhage (code I60.*). For 256 patients, aneurysmal subarachnoid hemorrhage on initial computed tomography (CT) scan was verified by an experienced neuroradiologist (H.W.). In a further step, 18 (7.0%) patients who did not receive aneurysm repair because of disease severity were excluded, resulting in 238 patients as final study cohort (Fig. 1, Supplemental digital content (SDC) 1: Table S1 showing reasons for exclusion).

**FIGURE 1.**
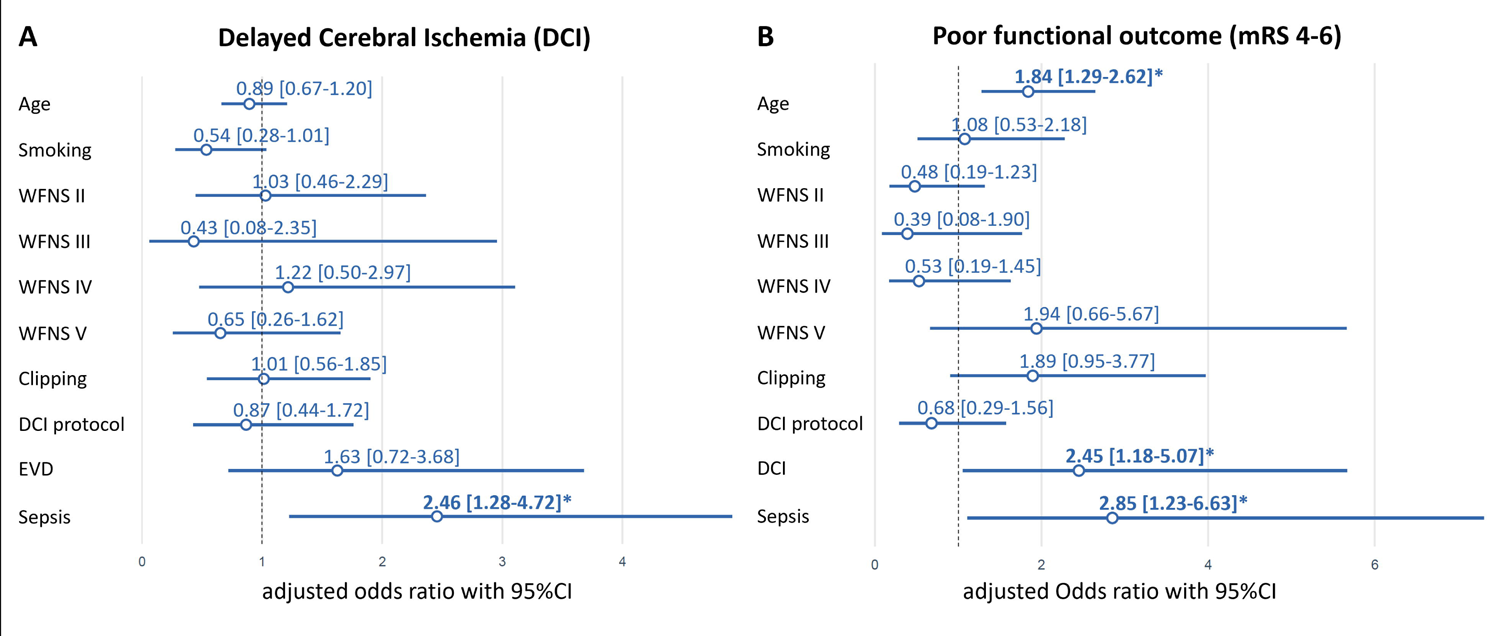
Flow diagram of study cohort selection. ICD indicates international classification of diseases; ICU, intensive care unit, SAH, subarachnoid hemorrhage.

### Clinical assessment

All patients were screened for healthcare-associated infections according to center for disease control (CDC) criteria^16^. For all patients who developed an infection, sequential organ failure assessment (SOFA) scores^17^ were determined for 24-hour periods by intensive care physicians. To ensure comparability of sepsis criteria application with the original approach of sepsis-3 definition^15^, sepsis-3 criteria were applied using an automated algorithm as previously described^18^. Baseline characteristics (age, sex, length of stay (LOS) in ICU and in-hospital mortality) were extracted from the electronic health records. Radiological imaging findings were evaluated by an experienced neuroradiologist (H.W.). Clinical parameters were extracted and verified by intensive care physicians: pre-existing illnesses, World Federation of Neurological Surgeons SAH grading scale (WFNS) at admission, placement of an external ventricular drainage (EVD), type of aneurysm repair and modified Rankin Scale (mRS) at hospital discharge. DCI as primary endpoint was defined as proposed by Vergouwen et al.^6^, based on neuroimaging of cerebral infarction from DCI with the following conditions: the presence of cerebral infarction on CT or MR scan of the brain within 6 weeks after SAH, or on the latest CT or MR scan made before death within 6 weeks, not present on the CT or MR scan between 24 and 48 hours after early aneurysm occlusion, and not attributable to other causes such as surgical clipping or endovascular treatment. Hypodensities on CT imaging resulting from ventricular catheter or intraparenchymal hematoma were not regarded as cerebral infarctions from DCI^6^. Clinical management was performed as described in detail previously and the procedures followed were in accordance with institutional guidelines^2,19^. Since January 2016, an updated protocol for standardized detection and management of DCI was implemented by the Department of Neurosurgery in line with our previous studies^19–21^. We adjusted for these changes in DCI treatment protocol during all our analyses by incorporating a binary (yes/no) factor variable (DCI protocol). Patients with infections and sepsis were treated in accordance with current recommendations^22^.

### Statistical analysis

Normally distributed variables were described using their means and standard deviations (SD), non-normally distributed variables with their medians, lower and upper quartile (LQ-UQ), while proportions were shown for categorical variables. All analyses were performed with the R statistics program™ (v.4.3.2, R Core Team 2023, Vienna Austria; RStudio IDE v. 2023.12.0 Build 554, Boston, MA, USA). Statistical analyses and their results were reported in accordance with the Statistical Analyses and Methods in the Published Literature (SAMPL) guidelines^23^.

The main research question, and explanatory variable of interest, was the effect of sepsis on the development of DCI. Therefore, we first performed propensity score matching (PSM) to adjust for potential confounders between subgroups of sepsis positive and negative patients^24^. Matching weights for sepsis-3 (n_yes_sepis-3_=55) subgroups were calculated using (the default settings of the matchit package) 1:1 nearest neighbor approach (N_total_=238; N_matched_=110) without replacement based on propensity score distance estimated with logistic regression including the following covariates: age, sex, WFNS grading scale, intraventricular hemorrhage (IVH), DCI protocol and smoking status as well as presence of arterial hypertension. The target estimand was calculated based on the average treatment effect of the treated (ATT). Second, we applied an exploratory modeling approach using standard generalized linear models (base glm) on the original cohort^25,26^. For this, the primary and secondary endpoint variables were the development of DCI (yes vs. no) and functional outcome at hospital discharge (mRS 0-3 vs. 4-6), respectively. These endpoints were modeled by multivariable logistic regression (LR) adjusted for sepsis, age, smoking status, WFNS grading scale, EVD-placement and type of aneurysm repair. The target explanatory variable of interest was sepsis based on sepsis-3 criteria. As sensitivity analyses, interaction terms were added to the multivariable LR model to investigate if the main- and combination effects of DCI and sepsis stayed consistent within mRS outcomes. Data regarding the time-dependency of these research questions were lacking in the literature. Thus, the distributions of time of occurrences of sepsis and DCI were described using median, LQ-UQ. Between group comparisons of un- and matched PSM cohorts (Table 2) were performed using Chi-squared test for categorical variables (with continuity correction) and two-group t-test. In case of potentially non-normally distributed continuous variables (e.g. ICU days), the Kruskal-Wallis (global test) or for two-groups the Wilcoxon-Mann-Whitney tests whereas for categorical variables with small cell counts the Fisher’s exact test were used^27,28^. Due to the limited number of cases, it was not possible to evaluate complex multivariable models in time-varying regression frameworks^29^. Figures were created using the ggplot2 grammar of graphics. P-values <0.05 were considered significant. Due to the explorative nature of our analyses, p-values were not adjusted for multiple testing^28^.

## Results

### Cohort characteristics

The 238 patients with aSAH had a median age of 56 years (Q1-Q3: 50-64, range:18-83; Table 1) and 70% were female. 116 (49%) patients developed any infection. When applying sepsis-3 criteria, 55 patients were diagnosed as septic, resulting in a sepsis frequency of 47% (55/116) in patients with an infection and 23% (55/238) in relation to the total cohort. This resulted in a mortality rate of 15% (8/55) within the septic subpopulation. Of all patients 31% (74/238) developed DCI, while 47% (26/55) with sepsis also developed DCI. The diagnosis of sepsis preceded the diagnosis of DCI in 65% (17/26) by a median of 43 hrs (mean=83 hrs, IQR= 49 hrs, range=1.85-468 hrs [19.5 days]). A total of 129 patients (54%) had a poor functional outcome at hospital discharge (mRS >3) and the overall in-hospital mortality was 15% (35/238, Table 1).

### Propensity score matching

PSM was performed for the development of sepsis. Variables included established risk factors for DCI^30^ such as smoking, sex, diabetes and arterial hypertension. Unmatched control cases were excluded from univariate analyses (n=128).

Before PSM (n=238, Table 2) diabetes, modified Fisher, IVH, EVD-placement, DCI, infection, functional outcome and ICU LOS were significantly different between septic and non-septic patients. After propensity score matching (n=110), infection (p<0.01) and ICU LOS (p=0.046) remained significant and among the 55 matched patients without sepsis, 27 (49%) had poor functional outcomes, contrasting with 43 patients (78%) who developed sepsis (p<0.01, Table 2).

### Association between sepsis and DCI

In multivariable LR fitted on complete cases (n=238, Table 1) of the original cohort, sepsis demonstrated an independent association with the occurrence of DCI (adjusted odds ratio (aOR)=2.46, 95%CI: 1.28-4.72, p=0.007; Table 3 and Fig. 2A). The history of smoking slightly missed statistical significance with DCI (p=0.054, Fig. 2A).

**FIGURE 2.**
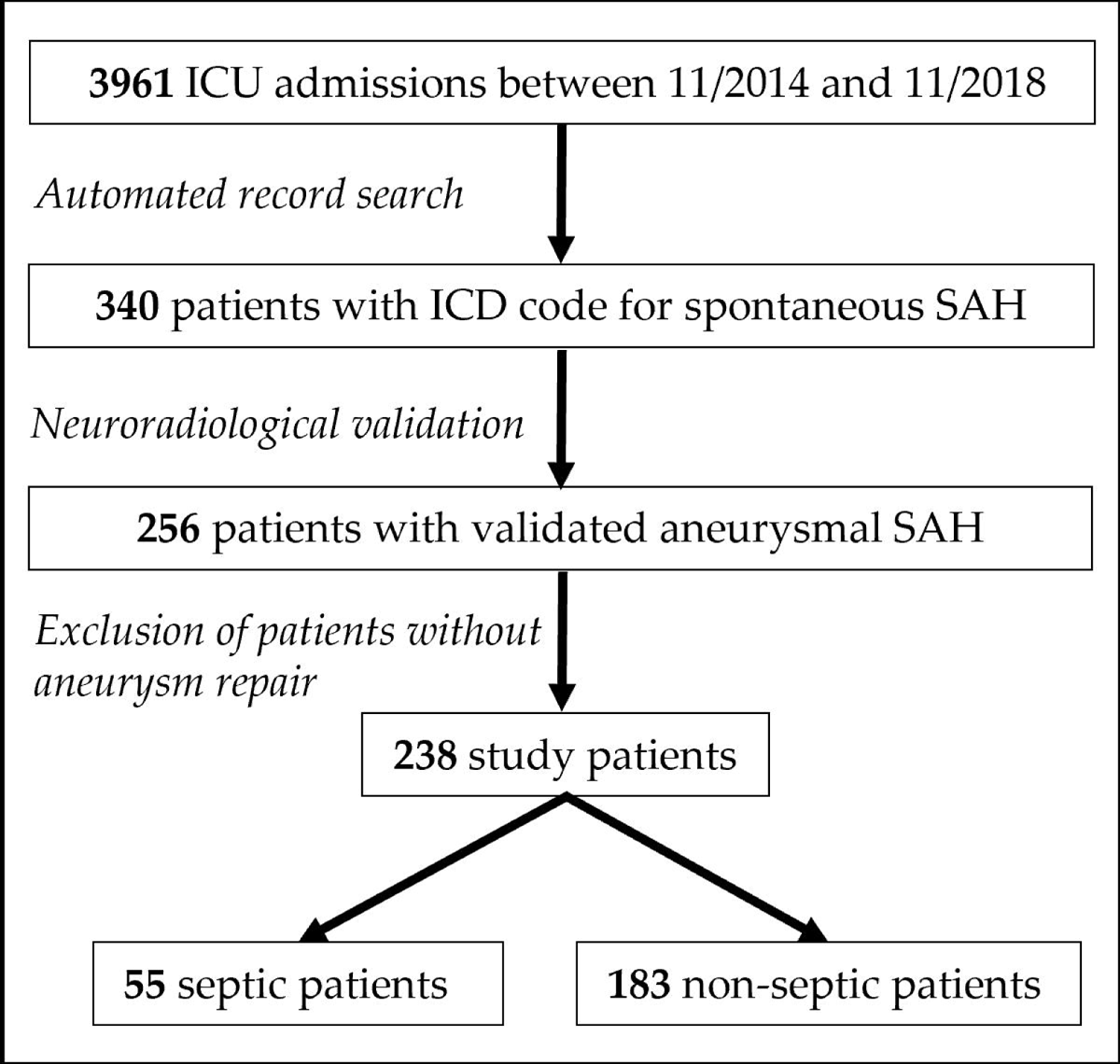
Multivariable logistic regression model-based adjusted odds ratios (aOR) for DCI (A) and poor (mRS 4-6) functional outcome (B). EVD for mRS 4-6 is not shown due to cleaner visibility and out of range point estimates and confidence intervals (aOR=21.87, 95%CI:6.76-70.77, p<0.01). Bold font and * indicate significance at p<0.05-level. DCI indicates delayed cerebral ischemia; EVD, external ventricular drainage; mRS, modified Rankin Scale; WFNS, World Federation of Neurological Surgeons SAH grading scale.

To further scrutinize the link between DCI and sepsis, we performed two sensitivity analyses. First, we excluded patients who developed sepsis after DCI (n=9) from the cohort, only focusing on cases in whom sepsis preceded DCI. This LR model indicated a trend towards increased odds of DCI if sepsis was diagnosed (aOR=1.59, 95%CI: 0.78-3.24, p=0.21; Table 3). Second, patients who developed sepsis before DCI (n=17) were excluded, focusing on cases in whom DCI preceded sepsis. In this LR analysis, there was no apparent association between DCI and sepsis (aOR=0.85, 95%CI: 0.37-1.95, p=0.70; SDC 1: Table S2 showing subgroup analysis).

### Association of sepsis and DCI with functional outcome

In multivariable LR models containing all cases (n=238) of the original cohort, sepsis was independently associated with poor functional outcome (mRS >3) at hospital discharge with an aOR=2.85 (95%CI: 1.23-6.63, p=0.02; Fig. 2B and SDC 1: Table S3 showing results of multivariable LR model for functional outcome). Age (aOR=1.84, 95%CI: 1.29-2.62, p<0.01), EVD-placement (aOR=21.87, 95%CI: 6.76-70.77, p<0.01) and DCI (aOR=2.45, 95%CI: 1.18-5.07, p=0.02) were additional predictors of poor functional outcome. Furthermore, we evaluated the effects of potential interactions between sepsis and DCI regarding functional outcome (SDC 1: Table S4 showing sensitivity analysis with interaction). Sepsis showed an interaction with DCI (aOR=11.05, 95%CI: 1.31-93.27, p=0.03) and neither of these variables remained independently associated (p=0.53 for sepsis and p=0.27 for DCI) in the interaction model (SDC 1: Table S4). Thus, indicating that the main effect of sepsis and DCI on functional outcome could not be independently interpreted from each other.

The proportion of patients with good functional outcome (mRS 0-3) gradually decreased in subgroups of patients from 56% (76/135) in those with neither sepsis nor DCI, to 43% (21/48) in those with no sepsis but DCI, to 34% (10/29) in patients with sepsis but no DCI and to 8% (2/26) in those with both sepsis and DCI.

## Discussion

The primary aim of the present study was to investigate the association of sepsis with DCI and a potential cumulative effect of sepsis and DCI on functional outcome in a cohort of patients with aSAH using current sepsis definition. Because of overlapping pathophysiological features, we hypothesized that sepsis and DCI might be associated and have a cumulative effect on poor functional outcome. In our cohort of aSAH patients, sepsis was independently associated with DCI. However, when patients that developed sepsis before or after DCI were excluded, this association did not remain. If sepsis preceded DCI, we found a trend towards a higher probability of developing DCI, which was probably limited by the sample size. Nonetheless, sepsis was confirmed as an independent risk factor for poor functional outcome in patients with aSAH. The worst functional outcome was observed in patients who suffered from both sepsis and DCI.

Robust data on the association of sepsis and DCI in patients with aSAH are limited and inconclusive^13^. A previous study by Foreman et al. described an independent association between nosocomial infection and DCI^13^. However, in their study no differentiation between localized and systemic infection was made^13^. Jeon et al. reported that sepsis was neither associated with DCI nor with mRS>3 at 3 months after SAH^14^. In a relatively small cohort of 98 aSAH patients, Budohosky et al. found a trend towards increased odds of DCI in patients with sepsis (OR=4.4, 95%CI: 0.08–25.2; p=0.095)^9^. Although the used sepsis operationalization was shown to be important in studies on sepsis-associated outcomes in critically ill patients in general^18,31^ and in patients with aSAH specifically^2,32^, neither of the aforementioned studies included details about the utilized sepsis definition. Additionally, both studies were performed before 2016^9,14^ and did not apply the updated sepsis-3 definition that focuses on septic organ dysfunction^15^. To our knowledge, this is the first study evaluating sepsis-3 criteria for the association between sepsis and DCI in patients with aSAH.

We observed a strong association between sepsis and DCI in the total cohort. When patients who developed sepsis after DCI were excluded from the analysis, thereby focusing on the effect of sepsis preceding DCI, the association between sepsis and DCI was no longer statistically significant but indicated a potential trend towards a positive association with DCI. Thus, we assume that a larger, potentially multi-centric cohort could improve statistical power and underscore their temporal association. This notion was also supported by the sensitivity analysis focusing on patients who developed sepsis after DCI, in which no relevant association between sepsis and DCI could be identified. These findings hint to a rather complex interplay between sepsis and DCI.

The observed association between sepsis and DCI in the present study might be explained by overlapping pathophysiological effects of sepsis in the brain and DCI. Both conditions are characterized by vascular dysfunction including dysregulation of cerebral vascular tone and impairment of vascular autoregulation^3,9,32^. While DCI can occur independently from cerebral vasospasm, there is still a strong association between angiographic vasospasm with neurological worsening, cerebral infarction, and poor outcome^5^. Simultaneously, in contrast to systemic vasodilation in early sepsis, vasoconstriction of the resistance arterioles in the brain occurs in patients with sepsis-associated encephalopathy^33^. Of note is the fact that the titration of the systemic circulation to a specific blood pressure target with vasopressors will not automatically restore cerebral perfusion in septic patients^34^. In recent years, again for both sepsis-associated encephalopathy and DCI, new evidence has been established that factors beyond a disturbed macrocirculation of cerebral blood vessels play an important role in the pathophysiology including microcirculatory disturbances due to endothelial dysfunction, neuroinflammation and microthrombosis with consecutive disruption of the blood-brain barrier and cellular injury^11,35–37^.

While DCI is defined as cerebral ischemia due to sequelae of SAH^6^, sepsis can cause cerebral ischemia via the multifactorial pathophysiological processes described above^7,8,35^. Hence, both sepsis and DCI are associated with derangements of vascular tone and integrity, microthrombosis formation and neuroinflammation resulting in cerebral ischemia^4,8,11,35^. These pathophysiological effects are concordant with the results of the present study in that both sepsis and DCI were independent risk factors for poor functional outcome in our cohort of aSAH patients. Sepsis is known to be independently associated with persistent new cognitive impairment and functional disability among survivors in general populations^38^ and might contribute to brain dysfunction after SAH as a second hit to an already vulnerable brain^1^. Accordingly, the relevance of sepsis on the functional outcome in patients with aSAH was shown before^1,39,40^ and confirmed in the present study. Interestingly, our analysis revealed a significant interaction between sepsis and DCI and showed that their effects on functional outcome could not be interpreted independently from each other. Furthermore, sensitivity analyses suggested that temporal characteristics of sepsis and DCI occurrence are of relevance. In line with this, we observed a gradual decrease of the proportion of patients with good functional outcome in the presence of DCI and sepsis. Only 8% of patients who suffered from sepsis and DCI achieved an mRS 0-3 outcome, which underlines a cumulative effect that warrants further research. In this context, it has been recently shown that systemic infection can increase intracranial inflammatory responses in patients with aSAH^12^, which might be a promising approach to further investigate the here established associations between sepsis, DCI and functional outcome. Further investigation into the temporal course of sepsis, the systemic immune response and the development of DCI could be helpful to better define the relationship^13^.

Our study has several limitations: First, the data are derived from a single-center retrospective cohort study. However, the retrospective nature permitted the inclusion of all available information independent from temporal occurrence and for the comprehensive description of infectious complications. Concurrently, the retrospective design prevented conclusions about cause–effect inferences.

In conclusion, the present study is, to our knowledge, the first to demonstrate a significant association between sepsis and DCI in aSAH patients and indicate the relevance of their temporal association. Sepsis was confirmed as an independent risk factor for poor functional outcome at hospital discharge. Furthermore, the co-occurrence of sepsis and DCI led to a cumulative effect towards poor functional outcome. Our analyses highlight the significant impact of sepsis on the disease course of aSAH patients suggesting a complex interplay between sepsis and DCI, which warrants further research on this topic.

## Data Availability

Study design and reporting adhered to the STROBE guidelines. All data produced in the present study are available upon reasonable request to the authors.

## Acknowledgements

We are grateful to Prof. Dr. Gabriel J.E. Rinkel for his valuable comments and helpful advice.

## Tables

**TABLE 1.** Clinical and treatment characteristics of the total cohort with aneurysmal subarachnoid hemorrhage (N=238).

**TABLE 2.** Clinical and treatment characteristics before and after propensity score matching (PSM) for cases with sepsis.

**TABLE 3.** Multivariable logistic regression models including sensitivity analysis for the primary endpoint: delayed cerebral ischemia (DCI).

Tables.docx

## Supplemental digital content 1

**TABLE S1.** Reasons for exclusion during neuroradiological validation.

**Table S2.** Sensitivity analysis: multivariable logistic regression model (N_sens_=221) for delayed cerebral ischemia (DCI) excluding patients who developed sepsis before DCI (N_excluded_=17).

**TABLE S3.** Multivariable logistic regression model for functional outcome (mRS 0-3 vs. 4-6).

**TABLE S4.** Sensitivity analysis including comparative statistics of multivariable logistic regression model (N=238) additionally adjusted for interaction between sepsis and DCI regarding functional outcome (mRS 0-3 vs. 4-6).

SDC1.docx

