## Supplemental Tables S1-S3 for "Association of sepsis and delayed cerebral ischemia in patients with aneurysmal subarachnoid hemorrhage"

**Supplemental digital content 1**

| TABLE S1. Reasons for exclusion during neuroradiological validation. | |
| --- | --- |
| Reason | **N** |
| Spontaneous SAH, no aneurysm | 14 |
| Traumatic SAH | 14 |
| Initial SAH not in study period, ICD-coding due to follow-up | 9 |
| No CT scan of initial SAH available | 6 |
| SAH initially treated elsewhere | 6 |
| Ischemic stroke with SAH component | 5 |
| Intracerebral bleeding with SAH component | 5 |
| No SAH | 3 |
| SAH already consolidated in initial CT | 2 |
| SAH associated with cerebral neoplasia | 2 |
| Arterio-venous malformation with SAH component | 2 |
| SAH as surgical complication | 2 |
| Subdural hematoma with SAH component | 1 |
| SAH associated with severe coagulation abnormalities due to |  |
| ECMO therapy | 5 |
| cardiac resuscitation | 3 |
| sepsis | 3 |
| cerebritis | 1 |
| Missing data | 1 |
| Total | 84 |
| CT indicates computed tomography; ECMO, extracorporeal membrane oxygenation; ICD, international classification of diseases; SAH, subarachnoid hemorrhage. | |

| TABLE S2. Sensitivity analysis: multivariable logistic regression model (N_sens_=221) for delayed cerebral ischemia (DCI) excluding patients who developed sepsis before DCI (N_excluded_=17). | |
| --- | --- |
| Variable | **aOR [95%CI]; p-value** |
| Age | 1.00 [0.97-1.02]; 0.85 |
| Smoking | 0.62 [0.32-1.19]; 0.15 |
| WFNS I-III vs. IV+V | 0.89 [0.46-1.69]; 0.71 |
| Clipping | 1.08 [0.59-2.00]; 0.80 |
| Sepsis | 0.85 [0.37-1.95]; 0.70 |
| To ensure statistical robustness, because only nine patients developed sepsis after DCI, the number of variables and their respective levels in the model were reduced. Thus, WFNS was dichotomized according to Abdulazim et al.^19^. aOR indicates adjusted odds ratio; DCI, delayed cerebral ischemia; EVD, external ventricular drainage; WFNS, World Federation of Neurological Surgeons SAH grading scale. | |

| TABLE S3. Multivariable logistic regression model for functional outcome (mRS 0-3 vs. 4-6). | |
| --- | --- |
| Variable | **aOR [95% CI]; p-value** |
| Age | **1.84 [1.29-2.62];** **<0.01** |
| Smoking | 1.08 [0.53-2.18]; 0.83 |
| WFNS grade II | 0.48 [0.19-1.23]; 0.13 |
| III | 0.39 [0.08-1.90]; 0.24 |
| IV | 0.53 [0.19-1.45]; 0.21 |
| V | 1.94 [0.66-5.67]; 0.23 |
| Clipping | 1.89 [0.95-3.77]; 0.07 |
| EVD placement | **21.87 [6.76-70.77]; <0.01** |
| DCI protocol | 0.68 [0.29-1.56]; 0.36 |
| DCI | **2.45 [1.18-5.07]; 0.02** |
| Sepsis | **2.85 [1.23-6.63]; 0.02** |
| aOR indicates adjusted odds ratio; DCI, delayed cerebral ischemia; EVD, external ventricular drainage; mRS, modified Rankin Scale; WFNS, World Federation of Neurological Surgeons SAH grading scale. Bold text indicates statistical significance at p<0.05 level. | |

| TABLE S4. Sensitivity analysis including comparative statistics of multivariable logistic regression model (N=238) additionally adjusted for interaction between sepsis and DCI regarding functional outcome (mRS 0-3 vs. 4-6). | |
| --- | --- |
| Variable | **aOR [95% CI]; p-value** |
| Intercept | **0.06 [0.02-0.23]; <0.01** |
| Age | **1.86 [1.29-2.67]; <0.01** |
| Smoking | 1.22 [0.59-2.49]; 0.59 |
| WFNS grade II | 0.55 [0.21-1.44]; 0.23 |
| III | 0.42 [0.08-2.13]; 0.30 |
| IV | 0.57 [0.21-1.60]; 0.29 |
| V | 2.32 [0.78-6.91]; 0.13 |
| Clipping | 2.01 [1.00-4.03]; 0.05 |
| EVD placement | **25.74 [7.39-89.59]; <0.01** |
| DCI protocol | 0.60 [0.25-1.41]; 0.24 |
| DCI | 1.58 [0.71-3.53]; 0.27 |
| Sepsis | 1.38 [0.51-3.72]; 0.53 |
| Interaction term (DCI*sepsis) | **11.05 [1.31-93.27]; 0.03** |
| *Model statistics* |  |
| Akaike information criterion (AIC) | 246.06 |
| Bayesian information criterion (BIC) | 291.20 |
| Pseudo R-squared | 0.49 |
| Bold p-values indicate statistical significance at the p<0.05 level.  aOR indicates adjusted odds ratio; DCI, delayed cerebral ischemia; EVD, external ventricular drainage; mRS, modified Rankin Scale; WFNS, World Federation of Neurological Surgeons SAH grading scale. | |
